# Multi-site Evaluation of SARS-CoV-2 Spike Mutation Detection Using a Multiplex Real-time RT-PCR Assay

**DOI:** 10.1101/2021.05.05.21254713

**Authors:** Carolin Bier, Anke Edelmann, Kathrin Theil, Rolf Schwarzer, Maria Deichner, Andre Gessner, Andreas Hiergeist, Ute Rentschler, Peter Gohl, Alison Kuchta, Chitra Manohar, Chris Santini, Dana Duncan, Jesse Canchola, Jingtao Sun, Gene Spier, Christian Simon

## Abstract

**Background:** SARS-CoV-2 causes COVID-19, which can be fatal and is responsible for a global pandemic. Variants with increased transmissibility or the potential to evade immunity have emerged and represent a threat to global pandemic control. Variants of concern (VOC) can be identified by sequencing of viral RNA, or by more rapid methods for detection of subsets of signature mutations.

**Methods:** We developed a multiplex, real-time RT-PCR assay (cobas^®^ SARS-CoV-2 Variant Set 1) for the qualitative detection and differentiation of three key SARS-CoV-2 mutations in the viral spike protein: del 69-70, E484K and N501Y. Analytical sensitivity and accuracy were evaluated at three testing sites using clinical specimens from patients infected with SARS-CoV-2 variants belonging to several different lineages, including B.1.1.7, B.1.351, and P.1.

**Results:** The limit of detection for E484K was between 180 and 620 IU/mL for the three different isolates tested. For N501Y, the LOD was between 270 and 720 IU/mL (five isolates), while for del 69-70, it was 80 - 92 IU/mL (two isolates). Valid test results were obtained with all clinical specimens that were positive using routine diagnostic tests. Compared to sequencing (Sanger and next-generation), test results were 100% concordant at all three loci; no false positive or false negative results were observed.

**Conclusions:** Data collected at three independent laboratories indicates excellent performance and concordance of cobas^®^ SARS-CoV-2 Variant Set 1 with sequencing. New sets of primers and probes that target additional loci can be rapidly deployed in response to the identification of other emerging variants.

## INTRODUCTION

SARS-CoV-2 is a novel coronavirus that causes COVID-19, a potentially lethal human respiratory disease [1]. SARS-CoV-2 has infected over 150 million individuals worldwide as of May 2021 [2] and is associated with more than 3.2 million deaths [2] and enormous economic impact across the world. SARS-CoV-2 is transmitted person-to-person via respiratory secretions, with clinical presentation varying from asymptomatic infection to mild illness to fatal disease [3-5]. The Coronaviridae family includes viruses that cause illness ranging from mild respiratory infection (human coronaviruses 229E, NL63, HKU1, and OC43) to more severe and often fatal diseases (MERS-CoV and SARS-CoV) [6].

Several lines of research and product development strategies are being pursued in order to combat the spread of SARS-CoV-2 and to mitigate the morbidity and mortality associated with COVID-19. These include the development of mRNA, virus vector, and protein subunit vaccines [7-11], small molecule antivirals [12, 13], immune modulators [14, 15], and other non-pharmaceutical interventions [16]. Vaccines that have been studied and developed to date are mostly focused on the viral envelope glycoprotein (Spike, S) and have been remarkably successful so far. In addition, monoclonal antibodies that target S have been developed as antivirals, and are effective treatments if administered soon after infection or symptom onset [17-19]. Monoclonal antibodies can also be administered to uninfected individuals to prevent infection by SARS-CoV-2 [20].

Like all RNA viruses, SARS-CoV-2 has a propensity to evolve in response to external selection pressures, due to an error-prone RNA-dependent RNA polymerase and large population sizes. While coronaviruses have a proof-reading function as part of the replicase complex, its high replication rate in each host and enormous population of infected people leads to the generation of a vast pool of viral variants from which more fit variants can emerge. Strong but incomplete inhibition of replication, which might occur in an infected person with partial immunity or treated with a single anti-S monoclonal antibody, is almost certain to result in the selection of SARS-CoV-2 variants with escape mutations in S that have higher replicative fitness than the wild-type virus in a population of susceptible hosts. Similarly, if a naturally occurring variant were to arise with increased ability to spread in an immunologically naïve population, it could out-compete the wild-type virus in a relatively short period of time. The emergence of several “ variants of concern” (VOC) in many different locations of the world in recent months is therefore not unexpected, and has several important public health and clinical implications [21-25]. Certain mutations in the S protein of these variants, such as a deletion of amino acids 69 and 70 in the N-terminal domain (NTD) and the N501Y substitution in the RBD, are characteristic features of specific VOC. Furthermore, some substitutions have been reported to confer altered susceptibility to neutralization by antibodies present following natural infection with the wild-type strain, elicited by vaccination, or by monoclonal antibodies used for treatment [26, 27]. Substitutions of unknown significance have also been reported, and more are likely to be detected in the future. Identification of VOC is considered to be essential to the public health response in some countries [28-30], as it may trigger specific actions related to containment efforts, such as duration of quarantine or isolation and re-testing frequency.

To monitor the spread of VOCs in the population, diagnostic tests that identify different mutations in the SARS-CoV-2 genome are needed. To date, variants have been characterized mainly by sequencing of the S gene or complete viral genomes, amplified by RT-PCR from patient specimens. Sequencing is sometimes preceded by putative VOC identification based on S-gene target amplification failure with standard primer sets [31]. Amplification and sequencing requires moderate to high viral loads, and requires resources that are not always readily available. An alternative approach is the use of assays that assess the presence of specific mutations, such as allele-specific PCR [32], high-resolution melting curve analysis [33], or other methods. These assays tend to have shorter turn-around time, greater sensitivity (i.e. require less starting material) and are less costly and labor-intensive compared to standard sequencing.

Here, we report the development and multi-site evaluation of a multiplex real-time PCR-based assay for SARS-CoV-2 variants with the E484K or N501Y substitutions or the 69-70 deletion in S. The analytical sensitivity of the assay was characterized using patient virus isolates whose titers were standardized to the First World Health Organization (WHO) International Standard for SARS-CoV-2 ribonucleic acid [34], and accuracy using clinical specimens was evaluated using a diverse panel of isolates from patients infected with wild-type or variant SARS-CoV-2.

## METHODS

### Assay design

The research use only (RUO) cobas^®^ SARS-CoV-2 Variant Set 1 (cobas SARSCoV2VarS1) is a single-well, multiplex, real-time RT-PCR test to be used with SARS-CoV-2 positive specimens. This assay is not for use in diagnostic procedures and is not CE marked or FDA cleared/approved. It detects the spike gene mutations del 69-70, E484K and N501Y in three different channels using three different fluorophores (JA270, FAM and HEX; Figure 1). Additionally the coumarin channel uses a single target in ORF1a/b region as a specimen check indicator (SCI) to re-confirm the presence and integrity of SARS-CoV-2 RNA. The CY5.5 channel serves as an internal control for the system process. The primers and probes were designed based on SARS-CoV-2 sequences in the GISAID and NCBI databases (total over 345,000 sequences) and were designed to provide maximum inclusivity for detecting the targets in each channel.

**Figure 1.**
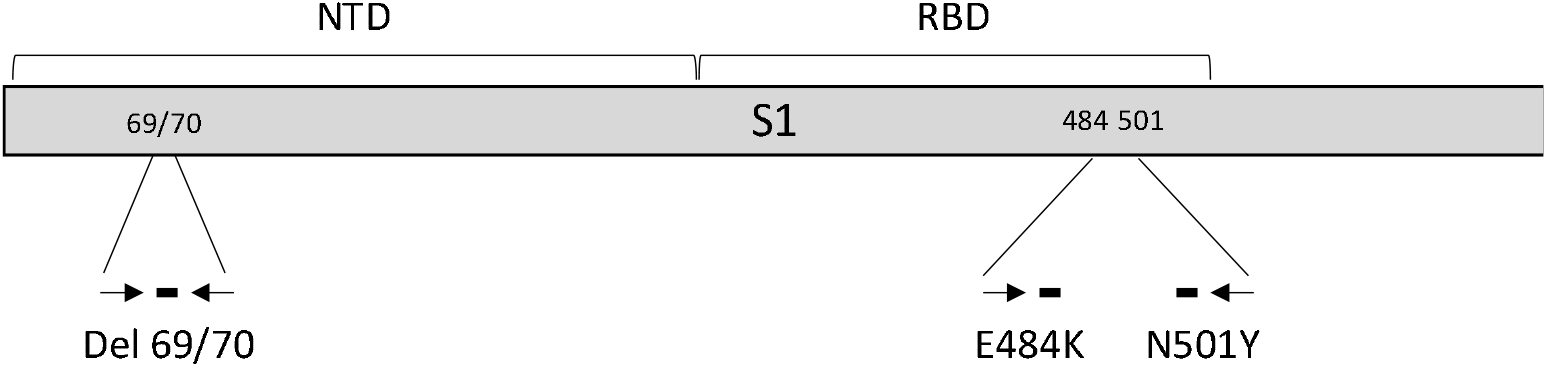
Illustration of the cobas® SARS-CoV-2 Variant Set 1 mutation detection assay design. NTD: N-terminal domain; RBD: receptor binding domain. The S1 portion (amino acids 1-686) of the spike gene is shown.

### Clinical specimens

Residual de-identified clinical specimens from three routine testing sites were used: Labor Berlin (Berlin, Germany), Bioscentia Labor (Ingelheim am Rhein, Germany), and the Institute of Clinical Microbiology and Hygiene, University Hospital of Regensburg (Regensburg, Germany). Patient specimens were collected and used according to institutional review board regulations in effect at each site.

Virus stocks for assay performance evaluation, including analytical sensitivity, were prepared by culturing virus from de-identified patient specimens in Vero cells.

### Testing sites

cobas SARSCoV2VarS1 testing and S gene sequencing were performed at three sites. Analytical sensitivity, analytical specificity, and accuracy studies were performed at Roche Diagnostics International AG (Rotkreuz, Switzerland). Analytical sensitivity and accuracy studies were performed at Labor Berlin. Accuracy studies were performed at University Hospital of Regensburg and Bioscentia, Ingelheim.

### Analytical sensitivity (limit of detection)

For determination of analytical sensitivity, six SARS-CoV-2 virus stocks grown in Vero cells in vitro were used. Two isolates each were prepared at the University of Zurich (isolate UZ1: P.2 lineage, clade 20B with E484K; isolate UZ2: B.1 lineage, clade 20A with N501Y), and at the University of Frankfurt (isolate UF1: B.1.351 lineage, clade 20H/501Y.V2 with E484K and N501Y; and isolate UF2: B.1.1.7 lineage, clade 20I/501Y.V1 with N501Y and del 69/70). Labor Berlin used two isolates which were kindly provided by the National Consultation Laboratory for Coronaviruses at the Institute of Virology, Medical University, Charité, Berlin (isolate LB1: B.1.351 lineage, clade 20H/501Y.V2 with E484K and N501Y; and isolate LB2: B.1.1.7 lineage, clade 20I/501Y.V1 with N501Y and del 69/70). cobas SARSCoV2VarS1 tests for the specimens from the University of Zurich and University of Frankfurt were performed at Roche Diagnostics International AG.

The titers of virus stocks were determined using the cobas® SARS-CoV-2 assay for use on the cobas® 6800/8800 system, which reports a cycle threshold (Ct) value. The First World Health Organization (WHO) International Standard for SARS-CoV-2 ribonucleic acid (RNA; NIBSC National Institute for Biological Standards and Control code 20/146) [34, 35] was also tested in this assay at two different concentrations at Roche Diagnostics International (3.7 and 5.7 log IU/ml), allowing conversion of Ct for unknown specimens to international units (IU) based on the linear regression of the standard curve (log_10_ IU/mL = 12.66 – 0.297*Ct).

Four to seven dilutions of each of the six different virus isolates were prepared in CPM (cobas® PCR Media) or a UTM-based simulated matrix (UTM, 50,000 human peripheral blood monocytes cells/mL, 0.05% mucin) to generate panels that included at least four concentrations: ∼3-fold, equal to, 0.3-fold, and 0.1-fold the expected limit of detection (LOD). Each panel member was tested in 21 replicates. LOD was determined using hit rate analysis (the concentration yielding at least 95% positive results) and reported in IU/mL.

### Accuracy using clinical specimens (agreement with sequencing)

To determine accuracy of the SARSCoV2VarS1 test, specimens containing SARS-CoV-2 with or without one or more of the three target loci were tested at three sites. The presence or absence of mutations was established by sequencing of S using next-generation methods (see Supplemental Methods). A total of 258 isolates were included. All specimens were RT-PCR positive using a variety of commercial or laboratory-developed tests. Several different specimen types including nasal, nasopharyngeal and oropharyngeal swabs, broncheo-alveolar lavage, tracheal secretions, and respiratory wash in diverse media (water, saline, universal transport medium, cobas PCR medium, etc.) were included (Table 1).

**Table 1.** Specimens used for accuracy evaluation

| Specimen type | Specimen collection site |  |  | Total |
| --- | --- | --- | --- | --- |
|  | Labor Berlin | University Hospital Regensburg | Bioscentia Ingelheim |  |
| Nasal, nasopharyngeal, or oropharyngeal swab | 148 | 16 | 44 | 208 |
| Throat irrigation fluid |  | 40 |  | 40 |
| Tracheal secretion |  | 7 |  | 7 |
| Broncheo-alveolar lavage |  | 2 |  | 2 |
| Processed sputum |  | 1 |  | 1 |
| **Total** | 148 | 66 | 44 | 258 |

### Analytical specificity (Interfering organisms)

Specificity was assessed using contrived specimens containing one of 17 different viruses (target concentration: 10^5^ units per mL in UTM-based simulated matrix), eight bacteria (10^6^ units per mL) or pneumocystis jirovecii (10^6^ units per mL). The 17 viruses tested were adenovirus, enterovirus, human coronavirus 229E, HKU1, NL63, and OC43, human metapneumovirus, influenza A and B virus, MERS-coronavirus, Parainfluenza virus 1, 2, 3 and 4, respiratory syncytial virus, human rhinovirus, and SARS-coronavirus. The eight bacteria were bordetella pertussis, chlamydia pneumoniae, haemophilus influenzae, legionella pneumophila, mycobacterium tuberculosis, mycoplasma pneumoniae, streptococcus pyogenes, streptococcus pneumoniae.

## RESULTS

### Analytical sensitivity (limit of detection)

The lowest virus concentrations tested that gave at least 95% positive results for each locus, as well as the corresponding mean cycle threshold values for the SCI control, are shown in Table 2. The limit of detection (LOD) determined by this method for E484K was between 180 and 620 IU/mL for the three different isolates tested. For N501Y, the LOD was between 270 and 720 IU/mL (five isolates), while for the deletion of codons 69 and 70, it was 80 or 92 IU/mL. The LOD for the SCI positive control target was between 18 and 80 IU/mL.

**Table 2.**
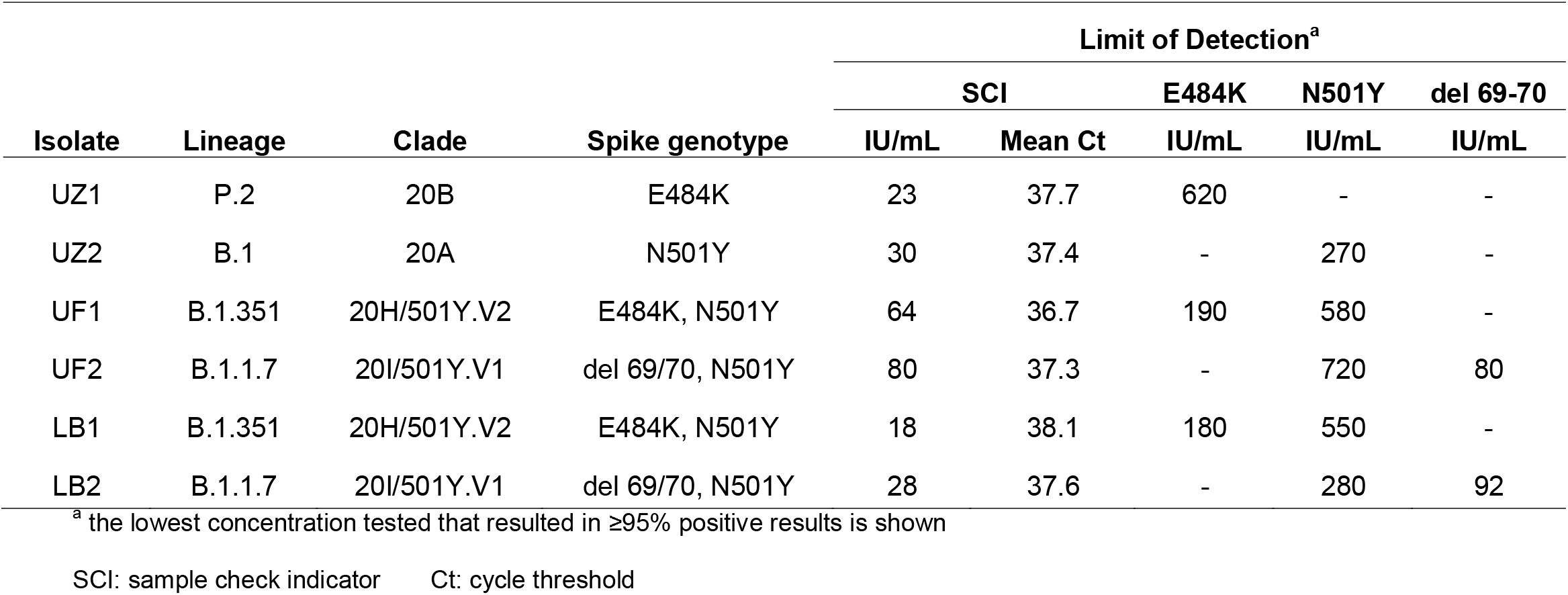
Assay Sensitivity (Limit of Detection)

| Isolate | Lineage | Clade | Spike genotype | Limit of Detection<sup>a</sup> |  |  |  |  |
| --- | --- | --- | --- | --- | --- | --- | --- | --- |
|  |  |  |  | SCI |  | E484K | N501Y | del 69-70 |
|  |  |  |  | IU/mL | Mean Ct | IU/mL | IU/mL | IU/mL |
| UZ1 | P.2 | 20B | E484K | 23 | 37.7 | 620 | - | - |
| UZ2 | B.1 | 20A | N501Y | 30 | 37.4 | - | 270 | - |
| UF1 | B.1.351 | 20H/501Y.V2 | E484K, N501Y | 64 | 36.7 | 190 | 580 | - |
| UF2 | B.1.1.7 | 20I/501Y.V1 | del 69/70, N501Y | 80 | 37.3 | - | 720 | 80 |
| LB1 | B.1.351 | 20H/501Y.V2 | E484K, N501Y | 18 | 38.1 | 180 | 550 | - |
| LB2 | B.1.1.7 | 20I/501Y.V1 | del 69/70, N501Y | 28 | 37.6 | - | 280 | 92 |

<sup>a</sup> the lowest concentration tested that resulted in ≥95% positive results is shown
SCI: sample check indicator Ct: cycle threshold

The differences we observed in LOD between targets could be a result of several factors, including primer and probe binding characteristics and local sequence context. The existence of such differences raise the potential for specimens with detectable RNA as evidenced by the SCI control and an undetectable result for E484K or N501Y that is due to the higher LOD for these targets, rather than absence of the mutation. The lowest SCI cycle threshold corresponding to the concentration at which ≥95% of replicates were positive for each of the three targets was 33.3 fo E484K, 33.4 for N501Y, and 36.0 for deletion of codons 69 and 70.

### Accuracy using clinical specimens (agreement with sequencing)

The SCI control reaction for all 258 isolates was positive, indicating the presence of viral RNA in the specimen. A total of 15 specimens with E484K present according to sequencing were tested (Table 3); all were reactive with the E484K probe (sensitivity: 100%). Conversely, reactivity with E484K was absent in 243 specimens with no substitution at position 484 (specificity: 100%). Similar results were obtained for N501Y (94 specimens with the substitution, 164 without) and the deletion of codons 69 and 70 (101 specimens with the deletion, 157 without) and are summarized in Table 3. No cobas SARSCoV2VarS1 false positive or false negative results were observed.

**Table 3.** Assay accuracy (agreement with sequencing)

| Target | Result | Mutation present | Mutation absent | Overall agreement % (95% CI) |
| --- | --- | --- | --- | --- |
| E484K | positive | 15 | 0 | 100 (98.6 – 100) |
|  | negative | 0 | 243 |  |
| N501Y | positive | 94 | 0 | 100 (98.6 – 100) |
|  | negative | 0 | 164 |  |
| Del 69-70 | positive | 101 | 0 | 100 (98.6 – 100) |
|  | negative | 0 | 157 |  |

### Analytical specificity (Cross reacting-organisms)

No signal was observed for the SCI or any of the targeted mutations with any of the specimens containing potentially cross-reacting organisms.

## DISCUSSION

The data presented in this report demonstrate that the newly developed cobas SARS-CoV-2 Variant Set 1 test is highly accurate for detection of SARS-CoV-2 variants containing the E484K or N501Y substitutions or deletion of codons 69 and 70. The presence of these mutations can be reliably detected when present at low concentrations, ranging from 80 to 720 IU/mL. No reactivity was observed with 26 different organisms that might be present in clinical specimens. Overall, these performance characteristics support the utility of this assay for surveillance for VOC, which may trigger public health actions. For example, information about the prevalence of VOC in the community or in specific locations may have implications for virus containment efforts and other public health actions such as contact tracing. Since E484K is associated with reduced susceptibility to bamlanivimab/LY-CoV555 (>100-fold in pseudovirus assays), casirivimab/REGN10933 (10-30-fold), C144-LS (>100-fold), and others [36-41], test results from population surveys may also inform public health decisions about the use of monoclonal antibodies for prevention or treatment in certain populations.

The LOD determined for this assay has been derived using material that is traceable to the WHO international quantitative standard, and reported in IU/mL. To date, sensitivity of SARS-CoV-2 RT-PCR tests has been reported using various non-standardized units including RNA copies/mL, TCID_50_/mL or pfu/mL. The use of international units will help improve standardization between platforms and testing sites, and allow more meaningful comparison and interpretations with regard to cutoffs to define virus detectability.

The use of real-time RT-PCR to detect variants with specific substitutions has several advantages compared to sequencing. An important one is the ability to generate results from specimens with low virus concentrations. The LOD reported here are 10 to 100-fold lower than the concentrations required to generate high quality sequence data (e.g. ∼10,000 copies/mL is usually needed for minimum sequencing depth using next-generation methods). Also, real-time RT-PCR variant detection can be performed more quickly, on more automated systems, and requires less intensive post-testing analysis. It is likely that overall testing costs will be lower, though this is dependent on details of the sequencing assay method and testing volume.

We note that the LOD for the E484K and N501Y substitutions is slightly higher than that of the SCI control signal (Table 2). Therefore, it is theoretically possible that a SARS-CoV-2 positive specimen that contains virus with the E484K or N501Y substitution could be reactive with the SCI control, but negative for these substitutions. Therefore, negative results for E484K and N501Y in specimens with Ct values for SCI above approximately 33 should be interpreted with caution.

This assay is not intended to replace diagnostic tests for SARS-CoV-2 infection, and has not been clinically validated for this purpose. Rather, the cobas SARSCoV2VarS1 assay should be performed as a follow-up test after confirmation of infection using an approved RT-PCR or antigen detection assay, in order to characterize the prevalence and spread of VOC in the population. Full sequencing is required in order to make an accurate assignment of each virus to a variant lineage or clade. In some contexts, point mutation assays could be used as part of a survey of variant prevalence.

Several different laboratory tests have been described [42-47], or are commercially available, which can help to identify specific mutations in the S gene of SARS-CoV-2. These tests are designed to detect certain VOC without the need for sequencing, and have varying sets of mutations targeted and other performance characteristics. The cobas SARSCoV2VarS1 assay is designed with several advantages compared to other tests, including full automation of nucleic acid extraction and real-time PCR reaction setup, internal controls, high throughput and short turn-around time (3.5 hours for 96 tests). Simplified workflow and short time to results are important features in the context of a rapidly evolving pandemic. It can be performed on the cobas 6800/8800 system [48], which can also be used for many other diagnostic applications in medium- to high-throughput clinical laboratories. The current set of primers and probes (Variant Set 1) provides information about the known substitutions at positions 484 and 501 and the deletion of codons 69 and 70, and thus provides information related to important VOC. Other substitutions in spike, including in the NTD, RBD and other regions, have also been reported in different VOC, some of which confer reduced susceptibility to one or more monoclonal antibodies (e.g. at positions 417, 453, 490, 494). Adaptation of the cobas SARSCoV2VarS1 assay, though development of additional sets of primers and probes, can be achieved readily when the need arises.

## Data Availability

Data are available on request to the authors

## Acknowledgements

We thank Dr. Trkola and Dr. Huber (Virology Department, University of Zurich, Switzerland), Dr. Ciesek (Virology Department, University of Frankfurt, Germany), and Victor Corman at Charité, Berlin for providing clinical specimens. The manuscript was prepared with assistance from Data First Consulting (Sebastopol, CA).

