## Supplemental Methods for "Multi-site Evaluation of SARS-CoV-2 Spike Mutation Detection Using a Multiplex Real-time RT-PCR Assay"

**Methods (Labor Berlin)**

Nucleic acids extraction was carried out on a MagNA Pure 96 system using 200 μl of sample material and the “Pathogen Universal Protocol” (elution volume, 50 μl).

For sequencing, RNA was reverse transcribed following the LunaScript RT SuperMix Kit (New England Biolabs) protocol.

Library preparation was done with the EasySeq RC-PCR SARS CoV-2 whole genome sequencing kit (Nimagen).

Typically, 94 libraries were multiplexed and sequenced in 2x 151 run mode on a NextSeq 500 system (Illumina) with a Mid Output kit v2.5 (300 cyles)

After sequencing, samples were demultiplexed using bcl2fastq v 2.20.0.422 with zero allowed barcode mismatches. Consensus genome sequences were generated with the Illumina workflow of the ncov2019-artic-nf pipeline (<https://github.com/connor-lab/ncov2019-artic-nf>), comprising the following steps: (a)  Read trimming using TrimGalore (<https://github.com/FelixKrueger/TrimGalore>) (b) Read mapping to the reference genome with Genbank accession MN908947.3 using bwa-mem (v0.7.18-r1188) and parameters -L 5,20, -O 5,12 -E 1 -B 5 (c) Read trimming of primer sequences and removal of mapped reads with length < 70bp or quality threshold < 20 using ivar (d) Consensus generation and variant calling using ivar with minimum frequency threshold of 0.9 and mimimum depth 20.

Afterwards, consensus sequences were manually curated to remove possible frameshifts introduced by alignment artifacts.
